# Informal coercion during childbirth: risk factors and prevalence estimates from a nationwide survey among women in Switzerland

**DOI:** 10.1101/2020.10.16.20212480

**Authors:** Stephan Oelhafen, Manuel Trachsel, Settimio Monteverde, Luigi Raio, Eva Cignacco Müller

## Abstract

**Background:** In many countries, the increase in facility births is accompanied by a high rate of obstetric interventions. Lower birthrates or elevated risk factors such as women’s higher age at childbirth and thus a higher need for control and security cannot entirely explain this rise in obstetric interventions. Another potential factor is that women feel coerced to agree to interventions; however, the prevalence of coercive interventions is unknown.

**Methods:** In a nationwide cross-sectional online survey, we assessed mothers’ satisfaction with childbirth and the prevalence of informal coercion during childbirth and of women at risk for postpartum depression. We used multivariable logistic regression to estimate the risk associated with multiple individual and contextual factors. Women at least 18 years old who gave birth in Switzerland within the previous 12 months were recruited online via Facebook ads or offline via various channels.

**Results:** A total of 6’054 women completed the questionnaire (drop-out rate 16.2%). An estimated 26.7% experience some form of informal coercion during childbirth. Having a cesarean section or instrumental vaginal birth was associated with an increased risk to experience informal coercion (all risk ratios > 1.5). The risk was also increased for women with a migrant background, women living in more urban regions and women with a risk pregnancy. Also, women to whom having a self-determined vaginal birth is important reported on informal coercion more often. Being at risk for postpartum depression was mostly associated with having an emergency cesarean section, having been transferred to hospital and the experience of informal coercion. Also, women with a migrant background seem to be at a higher risk to develop postpartum depression or having other mental health issues. Finally, women who had a non-instrumental vaginal birth reported higher satisfaction with childbirth experience and women who experienced informal coercion reported lower satisfaction.

**Conclusions:** One in four women experience informal coercion during childbirth, and this experience is associated with being at risk for postpartum depression and lower satisfaction with childbirth. Health care professionals should make every effort to prevent informal coercion and ensure sensitive aftercare for all new mothers in order to prevent traumatic effects.

## Background

When asked about their preferred way of giving birth, most women favor a vaginal birth with as few interventions as possible, including anesthesia [1-3]. This contrasts the high number of obstetric interventions in most middle- and high-income countries [4, 5]. Even for low-risk pregnancies, the rate of obstetric interventions during childbirth is rising. For example, large-scale studies from the U.S. and Canada indicate that around 60-90% of women who had planned to have a vaginal birth underwent one of the following interventions: induction of labor around term, epidural or spinal anesthesia, amniotomy, episiotomy, instrumental vaginal birth, or cesarean section (CS). Studies from Germany and the U.S. suggest that the observed increase in CS in recent decades is mainly attributable to rather subjective criteria or relative indications such as fetal distress or arrest of cervical dilation [6-8]. Other factors such as higher age of women at childbirth and the related increased rate of multiple births or obesity and associated risks cannot entirely explain the increase in CS [9].

Women’s preference for a safe birth could also be a factor contributing to the increase in obstetric interventions, even if the benefits of some interventions to improve fetal or maternal outcome are controversial [4, 10]. For example, in a Canadian survey involving 6421 women, 79.8% were satisfied with their overall birth experience, although the rate of obstetrical interventions was high [11]. In most regions of the world, the average fertility rate has dropped by more than 50% in the last 100 years, which may explain an increased need for safety and control during pregnancy and childbirth [7, 12]. Some women actually prefer a fast and ideally painless childbirth with interventions to a vaginal birth without interventions [2], especially if they feel anxious about giving birth or have had previous negative experiences [3, 5].

Given most women’s expressed preference for a vaginal birth and the social conditions affecting their preferences, it remains unclear to what extent the increase in obstetric interventions reflects their own safety concerns or medical indication. Importantly, these considerations also raise questions regarding the role of informal coercion when seeking women’s consent to interventions. Informal coercion denotes a range of measures on the continuum between self-determination and formal coercion, including inducement, persuasion, manipulation, pressure, or threats [cf. 13, 14-19]. In most jurisdictions, *formal* coercion during birth is only permissible under specific circumstances – i.e., when women lack decision-making capacity [20]. In psychiatry, the concept of informal coercion is advocated to prevent formal coercion [21, 22], such as forced medication or feeding. In obstetrics and gynecology, however, formal coercion is far less frequent, because women usually have decision-making capacity. Informal coercion may be more common in childbirth as an ultimate measure to urge women to accept diagnostic procedures or obstetric interventions. From an ethical and a legal point of view, these interventions are only admissible if women can accept or decline them freely, without undue influence or coercion, but with proper information and guidance from the HCP [23].

Research on the quality of maternity care conducted mostly in low-income countries has provided evidence for inadequate professional standards, including disrespectful, abusive or violent behaviors [2, 24-26]. In high-income countries, comparably subtle forms such as informal coercion might be more prevalent, given their health care systems’ emphasis on respecting patient autonomy and human rights [24]. Vedam et al. [27] report on women feeling coerced by HCP and that their physical complaints and needs are trivialized. In a representative study with 2400 U.S. women, about 15% of women who either had an induction of labor, epidural anesthesia, or CS felt pressured to accept the treatment [1]. About half of the women who wished to have a vaginal birth after a CS did not get this opportunity. Women who felt pressured had a doubled risk of labor induction and a sixfold risk of having a CS, even though there was no medical indication. In another cross-sectional study involving 2700 women from the U.S., 28% of women who gave birth in hospitals reported some form of mistreatment, most often unsupportive care, being shouted at or scolded, violation of privacy or being forced to accept treatment [24].

While conflicts between women and healthcare professionals (HCP) may be observable, informal coercion may come in covert forms, of which women or HCP may or may not be aware. HCP report on frequently pulling “the dead baby card”, where the mother is held responsible for a potential adverse outcome, irrespective of whether the baby is really at risk or not [28, 29]. Therefore, women’s reports on coercion depend on their level of knowledge about childbirth in general and about the specific rationale given for obstetric interventions [28, 30].

Due to the unknown extent of restrictions of women’s self-determination during childbirth, the goal of the current study was a) to assess how often and by what means informal coercion is experienced during childbirth, b) to evaluate how individual and contextual factors influence the risk for experiencing informal coercion and c) to understand to what degree this experience is associated with satisfaction with childbirth and postpartum depression.

## Methods

### Design

A nationwide cross-sectional survey was conducted among women who gave birth within the previous 12 months in Switzerland and were at least 18 years old. The recruitment phase lasted from August 2019 to January 2020. Respondents completed a self-administered questionnaire online.

### Recruitment

Following recommendations by Vehovar et al. [31], various recruitment procedures including both offline and online strategies were combined. Online, women were recruited via paid Facebook ads: They were redirected to the questionnaire by clicking on the Facebook ad. Offline, 180 pediatric or gynecological practices received ten leaflets each to distribute among women who met the inclusion criteria. The 180 practices were selected randomly in an online phone directory (https://tel.search.ch/). The number of selected medical practices in each Swiss canton reflected the number of births proportionally. However, the smaller French-speaking and Italian-speaking parts of Switzerland – compared to the German-speaking part – were slightly oversampled. By means of the newsletter of the Swiss Midwives Association, we asked midwives to distribute a direct link to the survey to women who met the criteria. Furthermore, an ad was posted in two Swiss parenting magazines. Women recruited via leaflets in medical practices and the parenting magazines had to enter a short vanity URL, which existed in four languages (i.e., German, French, Italian, and English; e.g., bfh.ch/birthstudy).

When designing recruitment material, neutral images/videos and vague wording were chosen to minimize selection bias [32], seeing as ad contents do not only influence the tendency for a subject to respond, but also impact Facebook’s ad delivery algorithms [33]. Both the leaflets and the Facebook ads were headlined “How did you experience the birth of your child?”. For the leaflets, an image of a newborn being held in someone’s hands was chosen (AdobeStock #193629437) and for the Facebook ad, a short video clip showing a close-up of a newborn on its mother’s chest (AdobeStock #120284636).

Recruitment on Facebook took place in four waves. Campaigns were either stratified by language regions (German-, French- and Italian-speaking parts) or by age category where the budgets for each stratum were defined to slightly oversample smaller strata as known from the most recent national census data [34]. After three recruitment waves, it became evident that participants with a migrant background were underrepresented in some age groups. Therefore, strata in the last wave were defined by the joint distribution of age category and residency status and the strata budget was increased accordingly for quotas which were underrepresented.

### Questionnaire development

Questionnaire development began with a literature review on questionnaires assessing childbirth experience, patient satisfaction, patient participation, autonomy and respect, informal coercion, patient consent, mistreatment and abuse during childbirth, and items were adapted for the present study [1, 13, 14, 16, 27, 35-40]. Because the goal was to achieve a low dropout rate, a rather short questionnaire was developed and possibly more “interesting” questions about the birth itself were placed at the beginning of the questionnaire [41]. Questions were worded in a way that was both medically precise and understandable for laypeople to minimize bias favoring women with a higher level of education. For example, to check for risk pregnancy, instead of presenting a long checklist of complications and diseases, we asked women if they had required medical treatment during pregnancy, either as an in- or outpatient.

A first draft of the questionnaire was reviewed by 19 experts (eight obstetricians, nine midwives, one clinical ethicist and one clinical psychologist), i.e., they rated each question on a 4-point Likert scale in terms of relevance and clarity and commented on issues or suggested changes. A second draft was then pilot tested with 20 mothers who completed the questionnaire and commented on the clarity of the questions and missing aspects. The questionnaire was then translated to French, Italian and English by professional translators. Afterwards, three lab members fluent in each target language reviewed all items. As a final step, all four language versions were compared to each other item by item to guarantee consistency over language versions.

The questionnaire was set up using Qualtrics(tm), with a focus on mobile phone compatibility. All answer options were presented in random order unless there was a natural order (e.g., mother’s age category). When asked about informal coercion, forced-choice questions instead of select-all-that-apply were chosen, which seems to provide more accurate responses with regard to undesirable events [42, 43]. Because we assumed that questions about informal coercion could trigger additional thoughts among respondents, multiple open questions were included, which again seems to increase response rates [41].

### Outcome variables

Informal coercion was operationalized following the Swiss Academy of Medical Sciences’ guidelines on coercive measures in medicine [44] and assessed on two levels. First, all respondents were asked if they had felt pressured to consent to any intervention (*general*; see Table 1). Second, women who had undergone either a CS, instrumental vaginal birth, induction of labor, episiotomy, or amniotomy, were asked six questions regarding that specific intervention. These questions covered aspects of informed consent, opposition to the intervention as well as intimidation and manipulation by HCP (*intervention specific*). This questionnaire design allowed both the assessment of the prevalence of informal coercion -regardless of whether women had undergone certain obstetric interventions - and a more precise evaluation of informal coercion in the context of specific interventions. Additionally, we assessed verbal violence with one item, i.e., “Did any health professional address you in an insulting or derogatory manner?” (*Insult*).

**Table 1:**
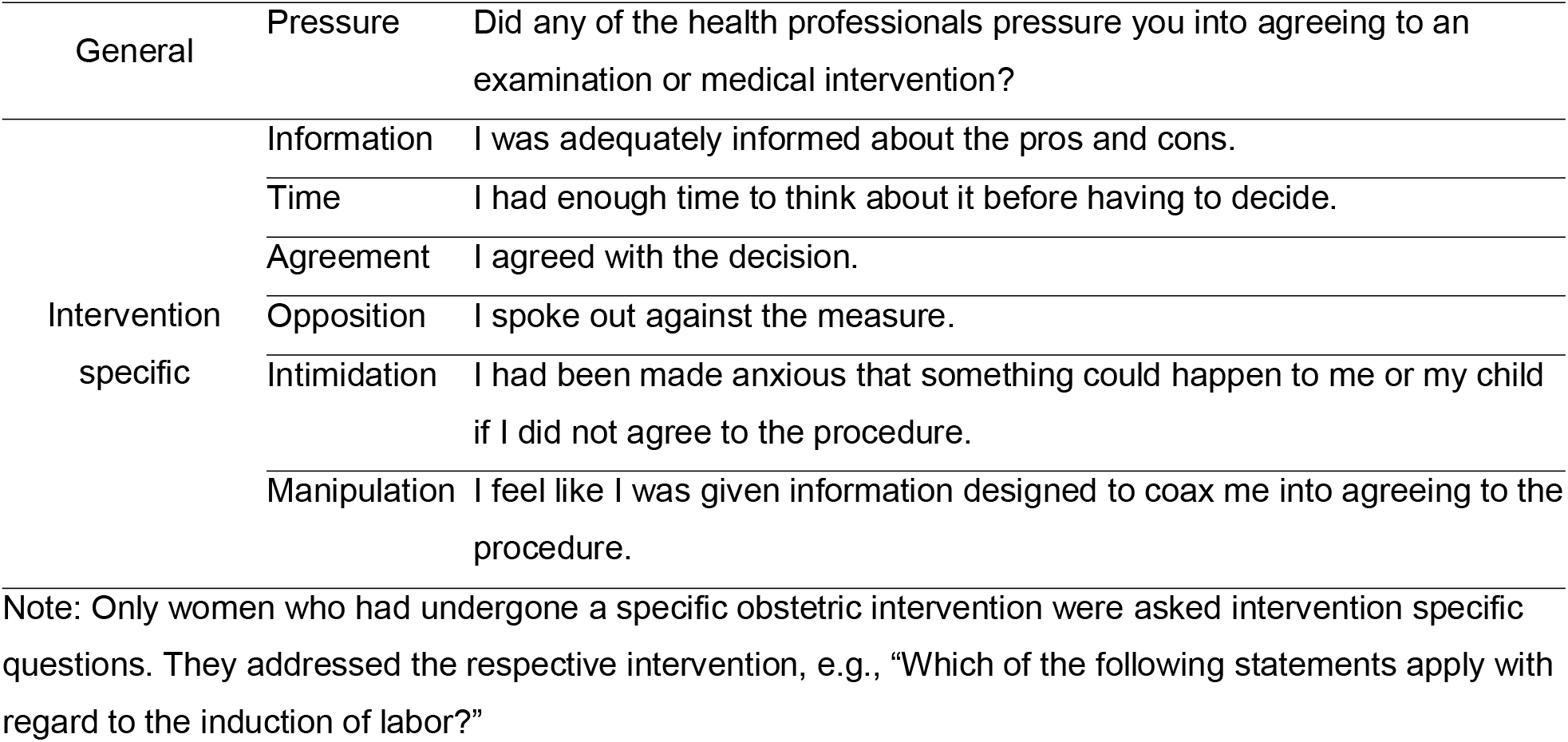
General and intervention specific items measuring informal coercion.

Initially, the criteria for informal coercion were considered fulfilled if at least one of the seven questions indicated that the woman had experienced informal coercion. However, due to various considerations, we then chose a more conservative and more reliable measure. First, there are numerous conceivable response patterns which do not imply coercion. For example, if a woman replies that she felt intimidated but at the same time felt well informed, agreed with the intervention, etc., it does not seem justified to assume coercion. It is also conceivable that, especially after hectic situations, women may state that they were neither sufficiently informed (*Information*) nor had enough time to reflect (*Time*). Again, these two items alone do not necessarily imply informal coercion. Therefore, respondents were categorized as having experienced informal coercion during childbirth if one or both of the following conditions applied: a) a respondent felt pressured to consent to medical interventions or diagnostics (*Pressure)*, b) two or more of the intervention specific items indicated a form of informal coercion, and at least one of these items was not *Information* or *Time*.

Satisfaction with childbirth experience was measured with the 12 items short version of Salmon’s item list [SIL; 45]. The SIL is a multidimensional instrument covering fulfilment, physical discomfort and emotional adaptation. While the original instructions state that respondents should reply regarding the whole birthing process including “the first hours after birth”, we asked women about their feelings which best describe their “childbirth experience”^1^.

To test a possible association between the experience of informal coercion and postpartum depression, we used the two validated Whooley questions [46]. The National Institute for Health and Care Excellence [47] recommends using them in the early postnatal period for depression identification. However, others suggest that they might lack specificity and indicate any mental health disorder, which then requires a more precise clinical assessment [48].

### Predictors and possible confounders

Based on the literature review mentioned above and the empirical findings outlined in the introduction, data regarding socio-demographical aspects, birth preparation, birth setting, birth history, pregnancy and birth characteristics, preference for active participation during childbirth, medical indications, medication, freedom of movement and other obstetric interventions were collected^2^. All variables are listed in Additional file 1: Table S1.

## Data analysis

The primary goal of the statistical analysis was to provide prevalence estimates of informal coercion. Therefore, procedures to weight the survey sample to represent all new mothers in Switzerland were applied accordingly. A raking algorithm was chosen based on the full joint distribution of age category and residence status and the marginal distributions of civil status, place of birth, mode of delivery, nulliparity, and geographical region, and subsequently trimmed weights larger than five times the average weight [49-51]. Weights were calculated based on the most recent census data available from the Swiss Federal Statistical Office [34, 52-54]. For non-hospital births, data provided by the Swiss interest group of birthing centers (IGGH-CH®) and the Swiss Association of Midwives were used [55, 56].

All analyses were conducted using R 3.6.3 [57]. Data were first imported using the qualtRics package [58]. Missing data of completed questionnaires were imputed using the mice package [59], using default imputation methods, with 50 imputations and a maximum of 30 iterations. Imputed datasets were then weighted using the rake procedure of the survey package [60]. Associated risk ratios (RR) were based on multivariable logistic regression models and on multivariable Poisson regression models in cases of a prevalence of ≥10% [61]. Overall, we included all possibly relevant predictors for the experience of informal coercion, satisfaction with birth and postpartum depression. To reduce the dimensionality of a few selected variables, we used optimal scaling procedures [62, 63] and subsequent principal component analysis. Variables with arbitrary numerical scales were standardized to ease comparability of predictors.

One researcher categorized information provided to “other” answer options or open-ended questions and discussed it with a second researcher in cases of ambiguity. Responses to open-ended questions were used to validate data in closed questions and to either correct or delete entries that were clearly erroneous. Comments provided in “other” responses were either allocated to existing response options or formed a separate category.

## Results

### Survey response

A total of 7663 women accessed the first survey page and provided informed consent. Most participants were recruited via Facebook (6625, 86.5%; other channels: 1026, 13.5%). Because their most recent birth lay more than 12 months back, 428 women (5.6%) were excluded from the final analysis. Sixteen responses (0.22%) were excluded after checking the comments, mainly because the birth did not take place in Switzerland or because they were duplicate entries. Out of the 7226 women who started the questionnaire and met all eligibility criteria, 6054 (83.8%) completed it. Regarding missing data in all completed questionnaires, one question (birth duration) had 10.4% missing data, six had less than 4% and all others had less than 1%.

### Demographic statistics

Table 2 shows descriptive statistics of selected demographic information and birth characteristics of both the survey and census data. The census data were used to weight the sample data for all subsequent analyses. Overall, the survey sample overrepresented Swiss women who had a non-instrumental vaginal delivery and did not give birth in a hospital. Descriptive statistics of additional sociodemographic variables as well as the pregnancy and birth characteristics of the survey sample are summarized in Additional file 2: Table S2.

**Table 2:**
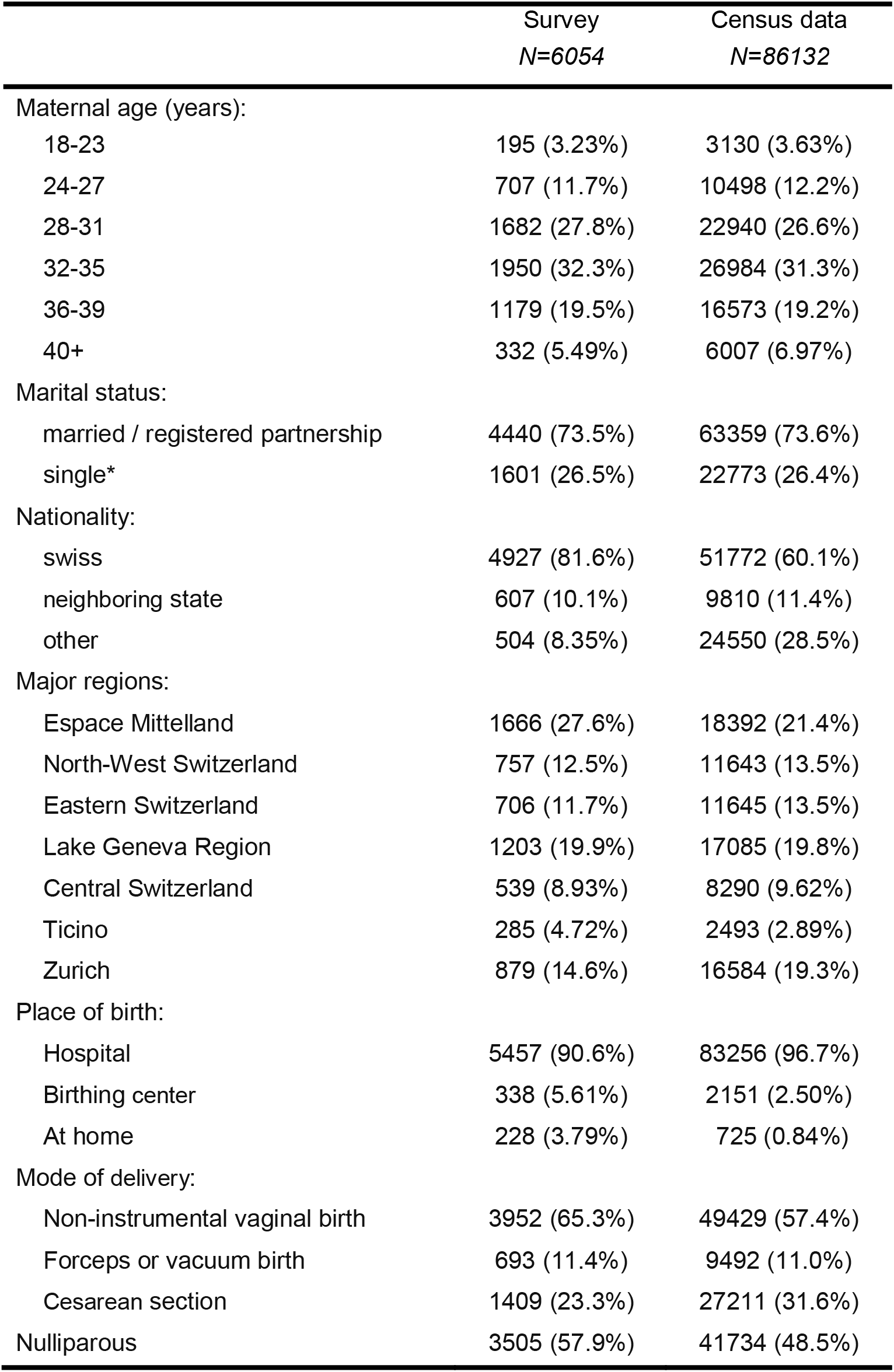
Descriptive statistics of final survey sample and census data for selected demographic and birth-related variables.

|  | Survey<br>*N=6054* | Census data<br>*N=86132* |
| --- | --- | --- |
| Maternal age (years): |  |  |
| 18-23 | 195 (3.23%) | 3130 (3.63%) |
| 24-27 | 707 (11.7%) | 10498 (12.2%) |
| 28-31 | 1682 (27.8%) | 22940 (26.6%) |
| 32-35 | 1950 (32.3%) | 26984 (31.3%) |
| 36-39 | 1179 (19.5%) | 16573 (19.2%) |
| 40+ | 332 (5.49%) | 6007 (6.97%) |
| Marital status: |  |  |
| married / registered partnership | 4440 (73.5%) | 63359 (73.6%) |
| single* | 1601 (26.5%) | 22773 (26.4%) |
| Nationality: |  |  |
| swiss | 4927 (81.6%) | 51772 (60.1%) |
| neighboring state | 607 (10.1%) | 9810 (11.4%) |
| other | 504 (8.35%) | 24550 (28.5%) |
| Major regions: |  |  |
| Espace Mittelland | 1666 (27.6%) | 18392 (21.4%) |
| North-West Switzerland | 757 (12.5%) | 11643 (13.5%) |
| Eastern Switzerland | 706 (11.7%) | 11645 (13.5%) |
| Lake Geneva Region | 1203 (19.9%) | 17085 (19.8%) |
| Central Switzerland | 539 (8.93%) | 8290 (9.62%) |
| Ticino | 285 (4.72%) | 2493 (2.89%) |
| Zurich | 879 (14.6%) | 16584 (19.3%) |
| Place of birth: |  |  |
| Hospital | 5457 (90.6%) | 83256 (96.7%) |
| Birthing center | 338 (5.61%) | 2151 (2.50%) |
| At home | 228 (3.79%) | 725 (0.84%) |
| Mode of delivery: |  |  |
| Non-instrumental vaginal birth | 3952 (65.3%) | 49429 (57.4%) |
| Forceps or vacuum birth | 693 (11.4%) | 9492 (11.0%) |
| Cesarean section | 1409 (23.3%) | 27211 (31.6%) |
| Nulliparous | 3505 (57.9%) | 41734 (48.5%) |

### Informed consent and informal coercion

Table 3 shows descriptive data of the three aspects of informed consent (*Information, Time, Agreement*) and the three forms of informal coercion (*Opposition, Intimidation, Manipulation*) for different delivery modes and selected interventions. Similar procedures or procedures that co-occur often showed a similar pattern of ratings of informed consent and informal coercion. Women who had a planned CS reported high levels of being informed adequately, having enough time to decide, and agreeing with the procedure. In comparison, women who underwent an unplanned CS or an induction of labor, which often co-occur, reported lower levels of information and time and the highest ratings of having opposed the procedure and felt manipulated. Emergency CS was associated with the highest rate of informal coercion overall; 37% of the women who had an emergency CS felt intimidated. Instrumental birth and episiotomy had the lowest ratings of informed consent. For both interventions only about 30% of the women felt adequately informed. Only roughly 20% of the women who had an instrumental birth and 17% of the women who had an episiotomy felt they had enough time to make their decision. Finally, only about half of the women who had an amniotomy received adequate information about the procedure and had enough time to decide.

**Table 3:** Number and percentage of women reporting fulfilled informed consent requirements and experiences of informal coercion.

|  | Aspects of informed consent |  |  | Forms of informal coercion |  |  |
| --- | --- | --- | --- | --- | --- | --- |
|  | Information | Time | Agreement | Opposition | Intimidation | Manipulation |
| Planned cesarean section | 427 (90.7%) | 427 (90.7%) | 451 (95.8%) | 35 (7.4%) | 142 (30.1%) | 25 (5.3%) |
| Unplanned cesarean section | 384 (76.2%) | 322 (63.9%) | 481 (95.4%) | 43 (8.5%) | 111 (22.0%) | 36 (7.1%) |
| Emergency cesarean section | 199 (58.5%) | 98 (28.8%) | 311 (91.5%) | 20 (5.9%) | 126 (37.1%) | 13 (3.8%) |
| Forceps or vacuum birth | 213 (30.7%) | 143 (20.6%) | 616 (88.9%) | 22 (3.2%) | 114 (16.5%) | 27 (3.9%) |
| Induction of labor | 1136 (74.2%) | 1122 (73.3%) | 1384 (90.5%) | 152 (9.9%) | 407 (26.6%) | 151 (9.9%) |
| Amniotomy | 713 (52.5%) | 754 (55.5%) | 1277 (94.0%) | 32 (2.4%) | 59 (4.3%) | 50 (3.7%) |
| Episiotomy | 208 (30.0%) | 119 (17.2%) | 530 (76.5%) | 45 (6.5%) | 89 (12.8%) | 38 (5.5%) |

Pairwise associations between the experience of informal coercion and medical indications as reported by the women can be found in Additional file 3: Table S3. Overall, women reported higher levels of informal coercion when they did not understand the reason for an intervention. Additionally, all other interventions such as fundal pressure, vaginal examinations and medication administration were associated with an increased risk of experiencing informal coercion. For women who had the opportunity to discuss the birth afterwards with involved HCP, on the other hand, the risk was lower.

Using imputed and weighted data, the estimated probability of experiencing any form of informal coercion was 26.7%. Being treated in a derogatory or insulting manner at least once was reported by 9.5% of women. Pressure to consent was reported by 16.3% of women. Risk ratios and 95% confidence intervals of factors influencing the experience of informal coercion are shown in table 4 (left column). Women from a non-neighboring state had and increased risk of experiencing informal coercion (RR 1.45), as did women living in more urban cantons (RR 1.16). Both preferring autonomous decisions during childbirth (RR 1.15) and preferring a vaginal birth (RR 1.15) increased the risk of experiencing informal coercion. Furthermore, women with risk pregnancies reported a higher rate of informal coercion. For women who gave birth at a birthing center, an independent birth facility run by midwives, the risk was three times lower (RR 0.35). Women who did not give birth where they had initially planned to – because they had to be transferred from a birthing center or a different hospital – had an increased risk (RR 1.47). Instrumental vaginal birth and all types of CS were associated with a higher risk of informal coercion (all RRs >1.5). Interestingly, women reported informal coercion more often if more time had elapsed since the birth (RR 1.17).

**Table 4:**
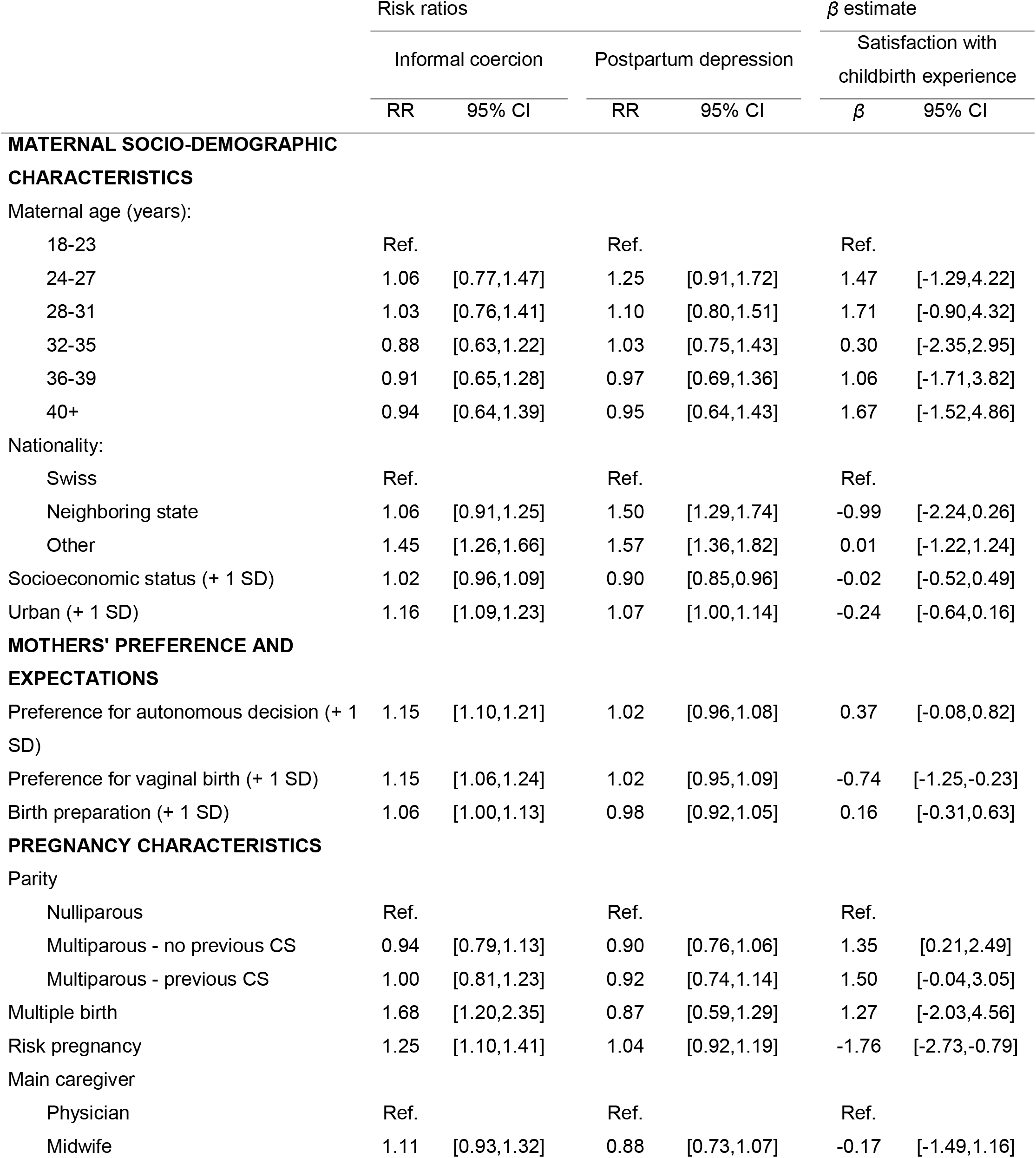
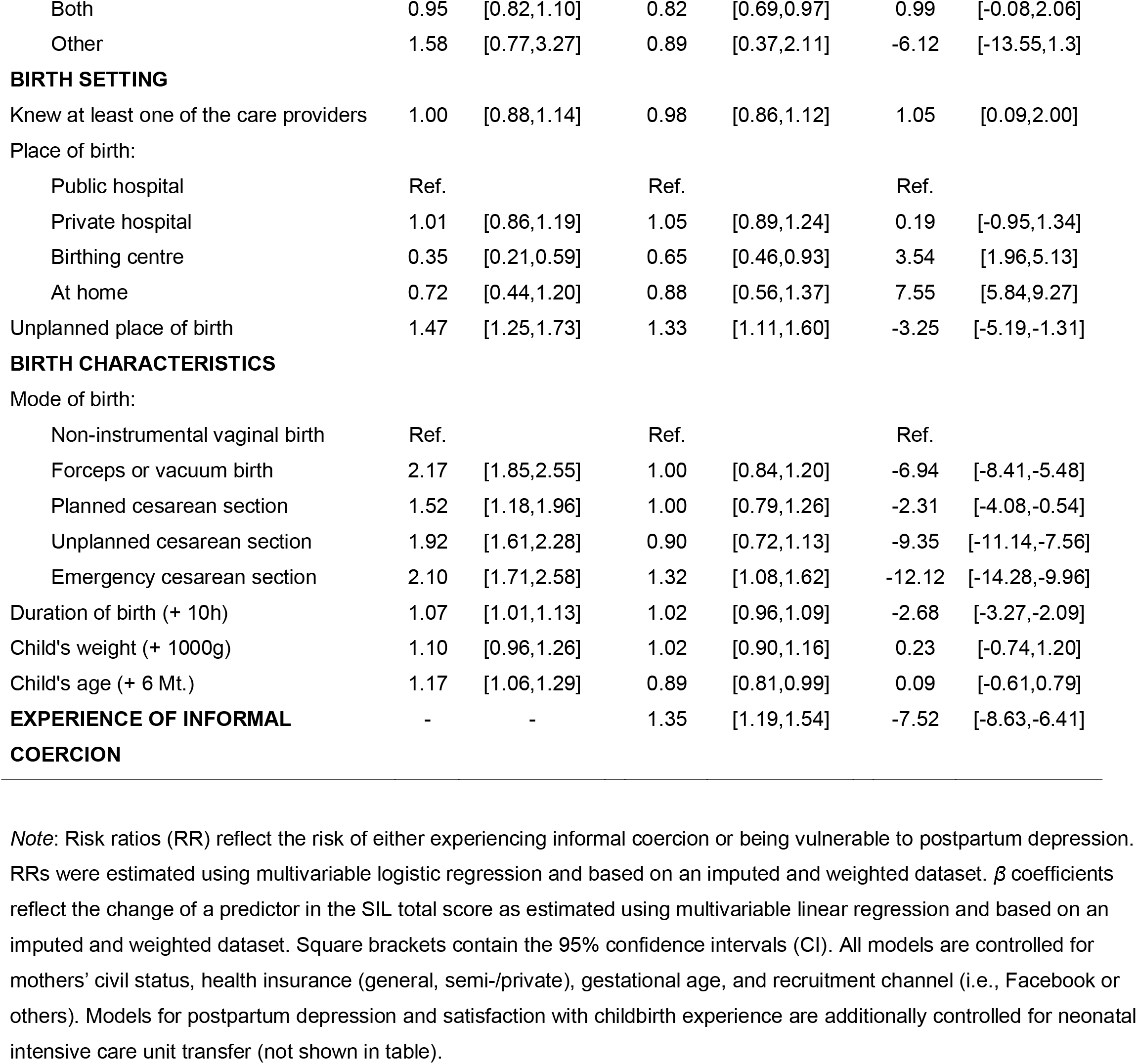
Estimated risks associated with experience of informal coercion, postpartum depression and satisfaction with childbirth experience.

| | Risk ratios | | | | $\beta$ estimate | |
| --- | --- | --- | --- | --- | --- | --- |
|  | Informal coercion |  | Postpartum depression |  | Satisfaction with childbirth experience |  |
| | RR | 95% CI | RR | 95% CI | $\beta$ | 95% CI |
| **MATERNAL SOCIO-DEMOGRAPHIC CHARACTERISTICS** |  |  |  |  |  |  |
| Maternal age (years): |  |  |  |  |  |  |
| 18-23 | Ref. |  | Ref. |  | Ref. |  |
| 24-27 | 1.06 | [0.77,1.47] | 1.25 | [0.91,1.72] | 1.47 | [-1.29,4.22] |
| 28-31 | 1.03 | [0.76,1.41] | 1.10 | [0.80,1.51] | 1.71 | [-0.90,4.32] |
| 32-35 | 0.88 | [0.63,1.22] | 1.03 | [0.75,1.43] | 0.30 | [-2.35,2.95] |
| 36-39 | 0.91 | [0.65,1.28] | 0.97 | [0.69,1.36] | 1.06 | [-1.71,3.82] |
| 40+ | 0.94 | [0.64,1.39] | 0.95 | [0.64,1.43] | 1.67 | [-1.52,4.86] |
| Nationality: |  |  |  |  |  |  |
| Swiss | Ref. |  | Ref. |  | Ref. |  |
| Neighboring state | 1.06 | [0.91,1.25] | 1.50 | [1.29,1.74] | -0.99 | [-2.24,0.26] |
| Other | 1.45 | [1.26,1.66] | 1.57 | [1.36,1.82] | 0.01 | [-1.22,1.24] |
| Socioeconomic status (+ 1 SD) | 1.02 | [0.96,1.09] | 0.90 | [0.85,0.96] | -0.02 | [-0.52,0.49] |
| Urban (+ 1 SD) | 1.16 | [1.09,1.23] | 1.07 | [1.00,1.14] | -0.24 | [-0.64,0.16] |
| **MOTHERS' PREFERENCE AND EXPECTATIONS** |  |  |  |  |  |  |
| Preference for autonomous decision (+ 1 SD) | 1.15 | [1.10,1.21] | 1.02 | [0.96,1.08] | 0.37 | [-0.08,0.82] |
| Preference for vaginal birth (+ 1 SD) | 1.15 | [1.06,1.24] | 1.02 | [0.95,1.09] | -0.74 | [-1.25,-0.23] |
| Birth preparation (+ 1 SD) | 1.06 | [1.00,1.13] | 0.98 | [0.92,1.05] | 0.16 | [-0.31,0.63] |
| **PREGNANCY CHARACTERISTICS** |  |  |  |  |  |  |
| Parity |  |  |  |  |  |  |
| Nulliparous | Ref. |  | Ref. |  | Ref. |  |
| Multiparous - no previous CS | 0.94 | [0.79,1.13] | 0.90 | [0.76,1.06] | 1.35 | [0.21,2.49] |
| Multiparous - previous CS | 1.00 | [0.81,1.23] | 0.92 | [0.74,1.14] | 1.50 | [-0.04,3.05] |
| Multiple birth | 1.68 | [1.20,2.35] | 0.87 | [0.59,1.29] | 1.27 | [-2.03,4.56] |
| Risk pregnancy | 1.25 | [1.10,1.41] | 1.04 | [0.92,1.19] | -1.76 | [-2.73,-0.79] |
| Main caregiver |  |  |  |  |  |  |
| Physician | Ref. |  | Ref. |  | Ref. |  |
| Midwife | 1.11 | [0.93,1.32] | 0.88 | [0.73,1.07] | -0.17 | [-1.49,1.16] |
| Both | 0.95 | [0.82,1.10] | 0.82 | [0.69,0.97] | 0.99 | [-0.08,2.06] |
| Other | 1.58 | [0.77,3.27] | 0.89 | [0.37,2.11] | -6.12 | [-13.55,1.3] |
| **BIRTH SETTING** |  |  |  |  |  |  |
| Knew at least one of the care providers | 1.00 | [0.88,1.14] | 0.98 | [0.86,1.12] | 1.05 | [0.09,2.00] |
| Place of birth: |  |  |  |  |  |  |
| Public hospital | Ref. |  | Ref. |  | Ref. |  |
| Private hospital | 1.01 | [0.86,1.19] | 1.05 | [0.89,1.24] | 0.19 | [-0.95,1.34] |
| Birthing centre | 0.35 | [0.21,0.59] | 0.65 | [0.46,0.93] | 3.54 | [1.96,5.13] |
| At home | 0.72 | [0.44,1.20] | 0.88 | [0.56,1.37] | 7.55 | [5.84,9.27] |
| Unplanned place of birth | 1.47 | [1.25,1.73] | 1.33 | [1.11,1.60] | -3.25 | [-5.19,-1.31] |
| **BIRTH CHARACTERISTICS** |  |  |  |  |  |  |
| Mode of birth: |  |  |  |  |  |  |
| Non-instrumental vaginal birth | Ref. |  | Ref. |  | Ref. |  |
| Forceps or vacuum birth | 2.17 | [1.85,2.55] | 1.00 | [0.84,1.20] | -6.94 | [-8.41,-5.48] |
| Planned cesarean section | 1.52 | [1.18,1.96] | 1.00 | [0.79,1.26] | -2.31 | [-4.08,-0.54] |
| Unplanned cesarean section | 1.92 | [1.61,2.28] | 0.90 | [0.72,1.13] | -9.35 | [-11.14,-7.56] |
| Emergency cesarean section | 2.10 | [1.71,2.58] | 1.32 | [1.08,1.62] | -12.12 | [-14.28,-9.96] |
| Duration of birth (+ 10h) | 1.07 | [1.01,1.13] | 1.02 | [0.96,1.09] | -2.68 | [-3.27,-2.09] |
| Child's weight (+ 1000g) | 1.10 | [0.96,1.26] | 1.02 | [0.90,1.16] | 0.23 | [-0.74,1.20] |
| Child's age (+ 6 Mt.) | 1.17 | [1.06,1.29] | 0.89 | [0.81,0.99] | 0.09 | [-0.61,0.79] |
| **EXPERIENCE OF INFORMAL COERCION** | - | - | 1.35 | [1.19,1.54] | -7.52 | [-8.63,-6.41] |

*Note:* Risk ratios (RR) reflect the risk of either experiencing informal coercion or being vulnerable to postpartum depression. RRs were estimated using multivariable logistic regression and based on an imputed and weighted dataset. $\beta$ coefficients reflect the change of a predictor in the SIL total score as estimated using multivariable linear regression and based on an imputed and weighted dataset. Square brackets contain the 95% confidence intervals (CI). All models are controlled for mothers' civil status, health insurance (general, semi-/private), gestational age, and recruitment channel (i.e., Facebook or others). Models for postpartum depression and satisfaction with childbirth experience are additionally controlled for neonatal intensive care unit transfer (not shown in table).

### Postpartum depression

The Whooley questions used for depression screening indicated that 27.0% of the women were at risk for postpartum depression or a different mental health disorder. Several demographic and birth-related factors were associated with an increased risk for possible mental health problems. Women living in urban cantons (RR 1.07) and women from other countries were more at risk (both RRs >1.5). Women who gave birth at a birthing center were less at risk (RR 0.65), while women who needed to be transferred had an increased risk (RR 1.33). Of all modes of delivery, only emergency CS was associated with an increased risk (RR 1.32). Experience of informal coercion also increased the risk for postpartum mental health disorders (RR 1.35).

### Satisfaction

Satisfaction with childbirth experience was measured with the total score of the SIL, with higher values indicating higher satisfaction. The main factors influencing satisfaction were place of birth, mode of delivery and the experience of informal coercion. Women who gave birth at home or at a birthing center were generally more satisfied with their birth than women who gave birth at a hospital (birthing center +3.54; at home +7.55). Women who did not give birth where they had planned to were less satisfied (−3.25). Women who had either an unplanned or an emergency CS had the lowest ratings of satisfaction (unplanned CS −9.35; emergency CS; −12.12). In addition, the experience of informal coercion had a negative effect on childbirth experience (−7.52).

## Discussion

The goal of the current study was to estimate the prevalence of informal coercion during childbirth in Switzerland and to assess the risk associated with individual and contextual factors. To do so, we developed a comprehensive questionnaire covering various aspects of informal coercion, satisfaction with childbirth experience, postpartum depression and a multitude of demographic, pregnancy and birth-related characteristics. Women at least 18 years old who gave birth in Switzerland within the previous 12 months were then recruited via different channels, although the majority filled in the questionnaire after clicking on a Facebook ad. An estimated 27% of women experienced informal coercion during childbirth. About 16% reported having felt pressured to consent to an intervention. In addition, the present data show that informal coercion negatively affects satisfaction with childbirth experience and is associated with an increased risk for postpartum depression.

Although the observed association between informal coercion and depression does not imply causality, it is worth noting that it is important to avoid informal coercion and enhance women’s childbirth experience as a means to prevent both the onset of postpartum depression and its exacerbation for women who already suffer from depression. Longitudinal studies suggest that having a CS may negatively affect women’s delivery experience and subsequently increase the risk for postpartum depression, especially for women who have a strong preference for vaginal delivery [64, 65]. Our data suggest that the relationship between mode of delivery and postpartum depression may be mediated by the experience of informal coercion.

Other studies report similar rates of informal coercion in high-income countries, although their scope and methodology differ. Vedam et al. [24] for instance found that about 28% of women who gave birth at a hospital experienced mistreatment. The most common forms of mistreatment were violations of physical privacy, being shouted at or scolded and unanswered requests for help. In another study, about 15% of women reported feeling pressured to consent to a medical intervention [1]. In general, the risk of experiencing mistreatment during childbirth seems to be lower if women are multiparous, older than 30 and white and if they speak the same language as the HCP. The risk is higher if women have to be transferred to hospital from a different place during childbirth [24, 66]. In line with previous research, the present study demonstrates that women with a migrant background were more at risk of experiencing informal coercion than Swiss nationals. The risk was also elevated for women living in more urban cantons compared to women living in more rural cantons. In urban cantons, both the established higher rate of CS [67] and the increased risk of informal coercion found in the present study suggest that a considerable number of interventions take place without explicit consent in urban areas, but this requires further exploration.

One important question in need of clarification is how the relationship between the birth setting and women’s preferences and expectations regarding childbirth impacts the experience of informal coercion. Our data shows that women who express a strong preference for a vaginal and self-determined birth tend to report informal coercion more often than women to whom this is less important. Expectations regarding a self-determined birth may be triggered by different conceptions of a “good” birth [68] and are not always realistic. In an institutional setting, women share the responsibility for their own health and that of their child with the HCP. Additionally, in an institutional setting, the birth process is subject to standardization for reasons of quality and effectiveness and HCP are required to follow specific guidelines. It may appear that birthing centers allow a more self-determined childbirth experience. However, a direct comparison between hospitals and birthing centers must be drawn with caution. First, only women with low-risk pregnancies can give birth at a birthing center. Second, birthing centers are not authorized to carry out most of the obstetric interventions requiring informed consent and bearing the risk of informal coercion outlined above. Lastly, the observed association between informal coercion and transfer to hospital does not necessarily imply any misconduct by HCP at the hospital. It could also reflect women’s disappointment that their wish for a vaginal birth with a minimum of medical interventions could not be met, even if they understand the reasons. Therefore, while the impact of women’s preferences and expectations on the experience of informal coercion may be established, the role of the birth setting and its relationship with women’s preferences and expectations requires further investigation.

In addition, the experience of informal coercion does not necessarily imply the HCP’s intention to exert coercion. Although working or organizational conditions can never justify the use of informal coercion, the following circumstances may explain why informal coercion can nevertheless occur and offer indications for its prevention. First, HCP may feel compelled to intervene immediately in case of doubt due to economic pressure, system-immanent incentives and fear of legal liability [18]. Second, most interventions undertaken by HCP are routine operations and HCP may not always have the sensitivity to consider an intervention’s possible consequences for the mother. Third, one must consider individual differences between people who perceive suggested measures as either ‘support’, a ‘nudge’ [69, 70], or outright pressure as well as the fact that this perception can change over time [71]. Fourth, HCP are confronted with a large variety of patient attitudes, preferences, and needs and some might not be able to deal with situations in which a woman rejects a treatment suggestion, even – or especially – if it is based on current best practice [29]. However, our findings suggest that informal coercion is a common feature of childbirth with potentially traumatizing consequences for the woman and it is likely to affect her family too. In general, obstetric interventions influence women’s birth experience negatively [56]. When weighing the possible beneficial and harmful outcomes of an intervention, HCP must also take into consideration that harmful outcomes include not only immediate physical, but also delayed and potentially longer-term psychological consequences for the mother.

In this study, about one in four women felt intimidated during childbirth. This number was even higher for women who had a CS or induction of labor (one in three). While many women prefer a vaginal birth, any concern for the child’s health tends to overrule other arguments when discussing possible interventions [72]. This is why “playing the dead baby card” is so effective, because none of the people involved – neither the woman nor the HCP – want to take responsibility for a negative outcome [73]. HCP have the power to modulate women’s fears and can either frighten them by stressing possible risks or empower them to take an active role in their childbirth. Of course, some women also have false beliefs due to a lack of knowledge of specific interventions. In the present study, women who did not understand the reasons for an intervention were more likely to feel coerced than women who did. Furthermore, women who were able to discuss the birth afterwards with involved HCP reported less coercion than women who did not have this opportunity. Both findings highlight the importance of the HCP’s duty to explain interventions and the reasons for them [1, 72]. Informing women about procedures and seeking their active participation is not only legally required, but also signals respect to the future mothers and their children.

### Strengths and limitations

To our knowledge, this is the first study to assess the prevalence of informal coercion in depth with a large-scale nationwide sample. Not only did we control for multiple pregnancy and birth characteristics but also for the women’s birth preparation and their attitudes and expectations regarding patient involvement. The questionnaire design ensured a comparably low dropout rate and therefore a representative sample.

One significant limitation is the possible self-selection bias which is usually more abundant in non-probability samples. On the one hand, the survey sample was not representative regarding variables such as place of birth, nationality, mode of delivery, and most likely also other characteristics that were not assessed. For example, the higher rate of women who gave birth at a birthing center, compared to census data, may be indicative of women who actively engage with the topic of childbirth and therefore are more interested in responding to a survey. Furthermore, while the recruitment material was carefully selected, there was no control of or insight into Facebook’s algorithms to deliver ads [33]. On the other hand, we followed several recommendations to decrease bias in non-probability samples, such as combining various channels for recruitment, both off- and online [31]. Moreover, women recruited via Facebook did not differ in reported experience of informal coercion, postpartum depression or satisfaction with childbirth from women recruited via other channels.

It is reasonable to assume that the reported prevalence is a rather conservative estimate. In general, when measuring patient satisfaction, more satisfied patients seem more inclined to respond to questionnaires than less satisfied patients [74]. For example, in our study, women who had a CS were more likely to drop out of the study if they did not want a CS, but not if a CS was their wish. Also, while we assessed informal coercion using multiple items, the rate of covert coercion – i.e., coercion that the women themselves did not recognize – is completely unknown. These micro-interactions are subtle and women who are overwhelmed and focused on the birth itself may not be aware of any interactions around them [75]. This feeling of being overwhelmed may continue for several months, which may explain why women reported less informal coercion in the first few months after birth than up to a year after.

## Conclusions

More than a quarter of women report that they experienced informal coercion during childbirth, i.e., that they did not agree with obstetric interventions or felt pressured or intimidated to consent. This is the case for all obstetric interventions, but even more so for unplanned CS, emergency CS, instrumental vaginal birth, and induction of labor. The experience of informal coercion is associated with an increased risk of postpartum depression. It is therefore essential to make every effort to prevent informal coercion. An increased focus on sensitive aftercare for all new mothers would allow HCP to detect women who experienced informal coercion and take measures necessary to prevent traumatic effects. In order to improve childbirth experience, a well-informed and empathetic debate on childbirth and on obstetric interventions and their consequences is necessary.

## Data Availability

The individual datasets generated during the present study are not publicly available, because the large variety of variables could allow de-anonymization of individual respondents. However, the final dataset used for analyses is available from the corresponding author on reasonable request.

## List of abbreviations

CS: Cesarean section
HCP: Healthcare professionals
RR: Risk ratio
SIL: Salmon’s item list

## Declarations

### Ethics approval and consent to participate

Because this was a nationwide study, all seven regional ethics committees in Switzerland confirmed that by the Swiss Human Research Act, the current study did not require ethical approval (Req-2019-00116). Participants were informed about anonymity and confidentiality of the study data. Informed consent was obtained on the first page of the questionnaire and respondents confirmed that they had understood the study information with a click.

### Consent for publication

Not applicable.

### Authors’ contributions

SO was responsible for study concept and design, and all other authors critically reviewed it. SO was responsible for statistical analyses and drafting the manuscript. All the authors were responsible for critical review and revision of the manuscript and contributed to data interpretations. SO and EC have full access to all the data in the study and assume responsibility for the integrity of the data and the accuracy of the data analysis. All authors read and approved the final manuscript.

## Acknowledgements

We would like to thank all the women who disclosed their intimate experiences during childbirth, Jacqueline Rusch for her valuable work developing the questionnaire and managing all technicalities to find an online solution, Stephanie Meyer for manuscript revision and content analysis, all experts and women who validated the questionnaire, the communications unit of Bern University of Applied Sciences for helping out with various recruitment issues, and Reto Bürgin for statistical support.

## Additional online files

Additional file 1: Table S1. List of independent variables, item wording, sources and possible transformations.

Additional file 2: Table S2. Descriptive statistics of additional sociodemographic variables, pregnancy, and birth characteristics of the survey sample.

Additional file 3: Table S3. Pairwise associations between the experience of informal coercion and medical indications, other interventions, and diagnostic procedures.

## Footnotes

1 Due to a programming error, the SIL items were initially not randomized, which was corrected after 29% of responses had been collected. An additional multivariable linear regression with the factor “SIL randomized yes/no” revealed no significant differences for predictor estimates.

2 Due to a wording error in the survey, it is likely that the frequencies for gestational age categories are not precise. The original item on gestational age differentiated between the categories, “< 32”, “32 – 36”, “37 – 41”, and “> 42” weeks. This made it impossible to correctly enter a gestational age of 42 weeks. The last category was adjusted to “> 41” after 65% of the data had been collected. However, this still leaves room for interpretation. Please note that gestational age did not significantly affect any of the outcomes, although we cannot exclude possible effects without this error.

## References

1. Declercq ER, Sakala C, Corry MP, Applebaum S, Herrlich A. Listening to MothersSM III: Pregnancy and Birth. New York: Childbirth Connection; 2013.

2. Downe S, Finlayson K, Oladapo O, Bonet M, Gülmezoglu AM. What matters to women during childbirth: A systematic qualitative review. PLOS ONE. 2018;13:e0194906. doi: 10.1371/journal.pone.0194906 PMID - 29664907.

3. Kringeland T, Daltveit AK, Møller A. What characterizes women who want to give birth as naturally as possible without painkillers or intervention? Sex Reprod Healthc. 2010;1:21–6. doi: 10.1016/j.srhc.2009.09.001 PMID - 21122592.

4. Miller S, Abalos E, Chamillard M, Ciapponi A, Colaci D, Comandé D, et al. Beyond too little, too late and too much, too soon: a pathway towards evidence-based, respectful maternity care worldwide. The Lancet. 2016;388:2176–92. doi: 10.1016/s0140-6736(16)31472-6 PMID - 27642019.

5. Boerma T, Ronsmans C, Melesse DY, Barros AJD, Barros FC, Juan L, et al. Global epidemiology of use of and disparities in caesarean sections. Lancet Lond Engl. 2018;392:1341–8. doi: 10.1016/s0140-6736(18)31928-7 PMID - 30322584.

6. Barber EL, Lundsberg LS, Belanger K, Pettker CM, Funai EF, Illuzzi JL. Indications Contributing to the Increasing Cesarean Delivery Rate. Obstetrics & Gynecology. 2011;118:29–38. doi: 10.1097/aog.0b013e31821e5f65 PMID - 21646928.

7. Bockenheimer-Lucius G. Zwischen „natürlicher Geburt”und „Wunschsectio”– Zum Problem der Selbstbestimmtheit in der Geburtshilfe. Ethik Med. 2002;14:186–200. doi: 10.1007/s00481-002-0189-y.

8. Metz TD, Stoddard GJ, Henry E, Jackson M, Holmgren C, Esplin S. How do good candidates for trial of labor after cesarean (TOLAC) who undergo elective repeat cesarean differ from those who choose TOLAC? American Journal of Obstetrics and Gynecology. 2013;208:458.e1-.e6. doi: 10.1016/j.ajog.2013.02.011 PMID - 23395923.

9. Lancet T. Stemming the global caesarean section epidemic. The Lancet. 2018;392:1279. doi: 10.1016/s0140-6736(18)32394-8 PMID - 30322560.

10. Rooks JP. Evidence-based practice and its application to childbirth care for low-risk women. Journal of Nurse-Midwifery. 1999;44:355–69. doi:10.1016/S0091-2182(99)00068-3.

11. Chalmers B, Dzakpasu S, Heaman M, Kaczorowski J. The Canadian Maternity Experiences Survey: An Overview of Findings. Journal of Obstetrics and Gynaecology Canada. 2008;30:217–28. doi: 10.1016/s1701-2163(16)32758-x PMID - 18364099.

12. Uphoff R. Aufklärung und Indikation zur Sectio als Beispiel für geburtshilflichen Paternalismus versus Geburtsmedizin als Dienstleistung für autonome Gebärende. 25 Jahre Arbeitsgemeinschaft - 25 Jahre Arzthaftung: Von der Krähentheorie bis zum groben Behandlungsfehler. Berlin, Heidelberg: Springer Berlin Heidelberg; 2011. p. 287–307.

13. Szmukler G, Appelbaum PS. Treatment pressures, leverage, coercion, and compulsion in mental health care. Journal of Mental Health. 2008;17:233–44. doi: 10.1080/09638230802052203.

14. Schweizerische Akademie der Medizinischen Wissenschaften (SAMW). Zwangsmassnahmen in der Medizin. Medizinisch-ethische Richtlinien der SAMW. Basel: SAMW; 2015.

15. Dondorp W, de Wert G. Prenatal Child Protection. Ethics of Pressure and Coercion in Prenatal Care for Addicted Pregnant Women. In: Hens K, Cutas D, Horstkötter D, editors. Parental Responsibility in the Context of Neuroscience and Genetics: Springer; 2017. p. 121–31.

16. Jäger M. Informeller Zwang in der therapeutischen Beziehung. Praxis. 2017;106:91–6. doi: 10.1024/1661-8157/a002585.

17. Valenti E, Banks C, Calcedo-Barba A, Bensimon CM, Hoffmann K-M, Pelto-Piri V, et al. Informal coercion in psychiatry: a focus group study of attitudes and experiences of mental health professionals in ten countries. Social Psychiatry and Psychiatric Epidemiology. 2015;50:1297–308. doi: 10.1007/s00127-015-1032-3 PMID - 25720809.

18. Kukura E. Obstetric Violence. Georgetown Law Journal. 2018;106:721–801.

19. Lorem GF, Hem MH, Molewijk B. Good coercion: Patients’ moral evaluation of coercion in mental health care. International Journal of Mental Health Nursing. 2015;24:231–40. doi:10.1111/inm.12106.

20. Glezer A. The Ethics of Court-Mandated Cesarean Sections. Journal of the American Academy of Psychiatry and the Law Online. 2018;46:276–8. doi: 10.29158/jaapl.003779-18.

21. Pelto-Piri V, Kjellin L, Hylén U, Valenti E, Priebe S. Different forms of informal coercion in psychiatry: a qualitative study. BMC Research Notes. 2019;12:787. doi: 10.1186/s13104-019-4823-x.

22. Elmer T, Rabenschlag F, Schori D, Zuaboni G, Kozel B, Jaeger S, et al. Informal coercion as a neglected form of communication in psychiatric settings in Germany and Switzerland. Psychiatry Research. 2018;262:400–6. doi: https://doi.org/10.1016/j.psychres.2017.09.014.

23. von Tigerstrom B. Informed Consent for Treatment: A Review of the Legal Requirements. Journal of Obstetrics and Gynaecology Canada. 2001;23:951–6. doi: 10.1016/S0849-5831(16)30863-1.

24. Vedam S, Stoll K, Taiwo TK, Rubashkin N, Cheyney M, Strauss N, et al. The Giving Voice to Mothers study: inequity and mistreatment during pregnancy and childbirth in the United States. Reproductive Health. 2019;16:77. doi: 10.1186/s12978-019-0729-2 PMID - 31182118.

25. Bohren MA, Vogel JP, Hunter EC, Lutsiv O, Makh SK, Souza JP, et al. The Mistreatment of Women during Childbirth in Health Facilities Globally: A Mixed-Methods Systematic Review. PLOS Medicine. 2015;12:e1001847. doi: 10.1371/journal.pmed.1001847.

26. Bowser D, Hill K. Exploring evidence for disrespect and abuse in facility-based childbirth. Boston: USAID-TRAction Project, Harvard School of Public Health.2010.

27. Vedam S, Stoll K, Rubashkin N, Martin K, Miller-Vedam Z, Hayes-Klein H, et al. The Mothers on Respect (MOR) index: measuring quality, safety, and human rights in childbirth. SSM - Population Health. 2017;3:201–10. doi: https://doi.org/10.1016/j.ssmph.2017.01.005.

28. Hall WA, Tomkinson J, Klein MC. Canadian Care Providers’ and Pregnant Women’s Approaches to Managing Birth: Minimizing Risk While Maximizing Integrity. Qualitative Health Research. 2011;22:575–86. doi: 10.1177/1049732311424292 PMID - 21940939.

29. Dexter SC, Windsor S, Watkinson SJ. Meeting the challenge of maternal choice in mode of delivery with vaginal birth after caesarean section: a medical, legal and ethical commentary. Bjog Int J Obstetrics Gynaecol. 2013;121:133–9; discussion 9-40. doi: 10.1111/1471-0528.12409 PMID - 24034671.

30. Sen G, Reddy B, Iyer A. Beyond measurement: the drivers of disrespect and abuse in obstetric care. Reproductive Health Matters. 2018;26:1–13. doi: 10.1080/09688080.2018.1508173 PMID - 30189791. 31.

31. Vehovar V, Toepoel V, Steinmetz S. Non-probability sampling. In: Wolf C, Joye D, Smith TW, editors. The Sage handbook of survey methods. London: SAGE Publications Ltd; 2016. p. 329–45.

32. Zhang B, Mildenberger M, Howe PD, Marlon J, Rosenthal SA, Leiserowitz A. Quota sampling using Facebook advertisements. Political Sci Res Methods. 2018:1–7. doi: 10.1017/psrm.2018.49.

33. Ali M, Sapiezynski P, Bogen M, Korolova A, Mislove A, Rieke A. Discrimination through optimization: How Facebook’s ad delivery can lead to skewed outcomes. 190402095. 2019. doi: 10.1145/3359301.

34. Federal Statistical Office. Lebendgeburten nach Alter der Mutter und Kanton, 1970-2019 [Live births by age of mother and canton, 1970-2019] 2020. https://www.bfs.admin.ch/bfs/de/home/statistiken/bevoelkerung/geburten-todesfaelle/geburten.assetdetail.13187380.html Accessed 25 Jun 2020.

35. Vedam S, Stoll K, Martin K, Rubashkin N, Partridge S, Thordarson D, et al. The Mother’s Autonomy in Decision Making (MADM) scale: Patient-led development and psychometric testing of a new instrument to evaluate experience of maternity care. PLOS ONE. 2017;12:e0171804. doi: 10.1371/journal.pone.0171804.

36. Scholl I, Kriston L, Härter M. PEF-FB-9–Fragebogen zur Partizipativen Entscheidungsfindung (revidierte 9-Item-Fassung). Klinische Diagnostik und Evaluation. 2011;4:46–9.

37. Birthrights. Dignity in Childbirth: the Dignity Survey 2013: Women’s and Midwives’ Experiences of Dignity in UK Maternity Care. Birthrights London; 2013.

38. Schrittenloher V. Peripartale Einflussgrößen auf Geburtmodus und Zufriedenheit unter besonderer Beachtung des Wunschkaiserschnittes: lmu; 2015.

39. Sjetne IS, Bjertnaes OA, Olsen RV, Iversen HH, Bukholm G. The Generic Short Patient Experiences Questionnaire (GS-PEQ): identification of core items from a survey in Norway. BMC Health Services Research. 2011;11:88. doi: 10.1186/1472-6963-11-88.

40. Dencker A, Taft C, Bergqvist L, Lilja H, Berg M. Childbirth experience questionnaire (CEQ): development and evaluation of a multidimensional instrument. BMC Pregnancy and Childbirth. 2010;10:81. doi: 10.1186/1471-2393-10-81 PMID - 21143961.

41. Burns KEA, Duffett M, Kho ME, Meade MO, Adhikari NKJ, Sinuff T, et al. A guide for the design and conduct of self-administered surveys of clinicians. Canadian Medical Association Journal. 2008;179:245–52. doi: 10.1503/cmaj.080372 PMID - 18663204.

42. Smyth JD, Christian LM, Dillman DA. Does “Yes or No” on the Telephone Mean the Same as “Check-All-That-Apply” on the Web? Public Opin Quart. 2008;72:103–13. doi: 10.1093/poq/nfn005.

43. Lau A, Kennedy C. When Online Survey Respondents Only ‘Select Some That Apply’. Washington DC: Pew Research Center. 2019. https://www.pewresearch.org/methods/2019/05/09/when-online-survey-respondents-only-select-some-that-apply/.

44. Swiss Academy of Medical Sciences (SAMS). Medical-ethical guidelines: Coercive measures in medicine. 2015.

45. Stadlmayr W, Bitzer J, Hösli I, Amsler F, Leupold J, Schwendke-Kliem A, et al. Birth as a multidimensional experience: comparison of the English-and German-language versions of Salmon’s Item List. Journal of Psychosomatic Obstetrics & Gynecology. 2009;22:205–14. doi: 10.3109/01674820109049975 PMID - 11840574.

46. Whooley MA, Avins AL, Miranda J, Browner WS. Case-finding instruments for depression. Two questions are as good as many. J Gen Intern Med. 1997;12:439–45. doi: 10.1046/j.1525-1497.1997.00076.x.

47. National Institute for Health and Care Excellence. Antenatal postnatal mental health: Clinical management and service guidance (NICE Clinical guideline No. 192). 2014.

48. Howard LM, Ryan EG, Trevillion K, Anderson F, Bick D, Bye A, et al. Accuracy of the Whooley questions and the Edinburgh Postnatal Depression Scale in identifying depression and other mental disorders in early pregnancy. The British Journal of Psychiatry. 2018;212:50–6. doi: 10.1192/bjp.2017.9 PMID - 29433610.

49. Kalton G, Flores-Cervantes I. Weighting Methods. Journal of official statistics. 2003;19:81–97.

50. Haziza D, Beaumont J-F. Construction of Weights in Surveys: A Review. Stat Sci. 2017;32:206–26. doi: 10.1214/16-sts608.

51. Lavrakas P. Encyclopedia of Survey Research Methods. 2011. doi: https://dx.doi.org/10.4135/9781412963947.

52. Federal Statistical Office. Lebendgeburten nach Staatsangehörigkeit und Alter der Mutter, 2000-2019 [Live births by nationality and age of mother, 2000-2019] 2020. https://www.bfs.admin.ch/bfs/de/home/statistiken/bevoelkerung/geburten-todesfaelle/lebenserwartung.assetdetail.13187373.html Accessed 25 Jun 2020.

53. Federal Statistical Office. Lebendgeburten nach Alter der Mutter und Geburtenfolge, 2005-2019 [Live births by age of mother and birth order, 2005-2019] 2020. https://www.bfs.admin.ch/bfs/de/home/statistiken/kataloge-datenbanken/tabellen.assetdetail.13187392.html Accessed 25 Jun 2020.

54. Federal Statistical Office. Lebendgeburten nach Geburtenfolge und Zivilstand der Mutter, 2005-2019 [Live births by birth order and marital status of mother, 2005-2019] 2020. https://www.bfs.admin.ch/bfs/de/home/statistiken/bevoelkerung/geburten-todesfaelle/geburten.assetdetail.13187451.html Accessed 25 Jun 2020.

55. Swiss interest group of birthing centers (IGGH-CH®). Statistik [Statistics] 2020. https://www.geburtshaus.ch/statistik.html Accessed 25 Jun 2020.

56. Swiss Association of Midwives. Statistikbericht der frei praktizierenden Hebammen der Schweiz [Statistical report of the freelance midwives in Switzerland] 2019. https://www.hebamme.ch/wp-content/uploads/2019/11/SHV_Statistikbericht_2019.pdf Accessed 25 Jun 2020.

57. R Core Team. Accessed R: A Language and Environment for Statistical Computing. Vienna, Austria: R Foundation for Statistical Computing; 2020.

58. Ginn J, Silge J. qualtRics: Download ‘Qualtrics’ Survey Data. R package version 3.1.2. 2020. https://CRAN.R-project.org/package=qualtRics.

59. Buuren Sv, Groothuis-Oudshoorn K. mice: Multivariate imputation by chained equations in R. Journal of statistical software. 2010:1–68.

60. Lumley T. Analysis of complex survey samples. R package version 3.33-2. 2019.

61. Barros AJ, Hirakata VN. Alternatives for logistic regression in cross-sectional studies: an empirical comparison of models that directly estimate the prevalence ratio. BMC Medical Research Methodology. 2003;3:21. doi: 10.1186/1471-2288-3-21.

62. Mair P, de Leeuw J. aspect: A General Framework for Multivariate Analysis with Optimal Scaling. R package version 1.0-5. 2018. https://CRAN.R-project.org/package=aspect.

63. Nguyen LH, Holmes S. Ten quick tips for effective dimensionality reduction. Plos Comput Biol. 2019;15:e1006907. doi: 10.1371/journal.pcbi.1006907 PMID - 31220072.

64. Eckerdal P, Georgakis MK, Kollia N, Wikström A-K, Högberg U, Skalkidou A. Delineating the association between mode of delivery and postpartum depression symptoms: a longitudinal study. Acta Obstetricia et Gynecologica Scandinavica. 2018;97:301–11. doi: 10.1111/aogs.13275.

65. Houston KA, Kaimal AJ, Nakagawa S, Gregorich SE, Yee LM, Kuppermann M. Mode of delivery and postpartum depression: the role of patient preferences. American Journal of Obstetrics and Gynecology. 2015;212:p229.e1-.e7. doi: https://doi.org/10.1016/j.ajog.2014.09.002.

66. Kukura E. Choice in birth: preserving access to VBAC. Penn State Law Review. 2009;114:955.

67. Federal Statistical Office. Anzahl und Rate der Kaiserschnitte nach Kanton und Wohnregion [Number and rate of cesarean sections by canton and region] 2019. https://www.bfs.admin.ch/bfs/de/home/statistiken/gesundheit/gesundheitswesen/spitaeler/patienten-hospitalisierungen.assetdetail.10787005.html Accessed 25 Jun 2020.

68. Karlström A, Nystedt A, Hildingsson I. The meaning of a very positive birth experience: focus groups discussions with women. BMC Pregnancy and Childbirth. 2015;15:251. doi: 10.1186/s12884-015-0683-0.

69. Gorin M, Joffe S, Dickert N, Halpern S. Justifying Clinical Nudges. Hastings Center Report. 2017;47:32–8. doi: 10.1002/hast.688.

70. Lantos JD. Ethical Problems in Decision Making in the Neonatal ICU. New England Journal of Medicine. 2018;379:1851–60. doi: 10.1056/NEJMra1801063.

71. Soltani H, Dickinson FM, Kalk J, Payne K. Breast feeding practices and views among diabetic women: A retrospective cohort study. Midwifery. 2008;24:471–9. doi: 10.1016/j.midw.2007.04.005 PMID - 17870219.

72. Coates R, Cupples G, Scamell A, McCourt C. Women’s experiences of induction of labour: qualitative systematic review and thematic synthesis. Midwifery. 2018;69:17–28. doi: 10.1016/j.midw.2018.10.013 PMID - 30390463.

73. Oelhafen S, Monteverde S, Cignacco E. Exploring moral problems and moral competences in midwifery: A qualitative study. Nursing Ethics. 2018:1–14. doi: 10.1177/0969733018761174.

74. French K. Methodological considerations in hospital patient opinion surveys. Int J Nurs Stud. 1981;18:7–32. doi: https://doi.org/10.1016/0020-7489(81)90004-3.

75. Darilek U. A Woman’s Right to Dignified, Respectful Healthcare During Childbirth: A Review of the Literature on Obstetric Mistreatment. Issues in Mental Health Nursing. 2018;39:538–41. doi: 10.1080/01612840.2017.1368752.

