## Additional file 1: Table S1 for "Informal coercion during childbirth: risk factors and prevalence estimates from a nationwide survey among women in Switzerland"

Additional File 1: Table S1. List of independent variables, item wording, sources and possible transformations.

| **Variable** | **Source / question** | **Response options / scale** | **Transformations / comments** |
| --- | --- | --- | --- |
| Birth preparation | How did you **prepare for the birth**? Multiple answers are possible. | - Antenatal classes - With books - Online - By writing a birth plan - No special preparation - Other:______ | Based on “other” responses, 9 new categories were built: *conversations* (with friends, HCP, etc.), *techniques* (hypnobirthing, meditation, visualization, etc.), *alternative therapies* (acupuncture, homeopathy, etc.), *information* (videos, apps, tv, social media), *yoga*, *sports*, *perineum preparation*, *diet* and *other*.  To reflect the amount of individual birth preparation, the sum of all categories selected by a respondent and a “yes” answer to the next question on considering various birth place options were entered in the model. |
|  | Did you consider various options before deciding on this place of birth? | - Yes - No |  |
| Main caregiver pregnancy | Which **specialist** was your main caregiver **during your pregnancy?** | - a doctor - a midwife - Other:______ |  |
| Risk pregnancy | Did you require medical treatment during your pregnancy? | - Yes, I was admitted to the hospital as an in-patient (overnight stay) - Yes, as an out-patient - No |  |
| Child’s age | How old is your child now? | 13 choices: “1 month” up to “12 months”, “more than 12 months” |  |
| Child’s weight | How much did your child weigh at birth approximately? | numerical value between 500 and 6000 grams |  |
| Gestational age | At how many **weeks of pregnancy** was your child born? | - < 32 (more than 8 weeks early) - 32-36 (more than 3 weeks early) - 37 - 41 - > 41 - I am not sure |  |
| Multiple birth | Was this a multiple birth? | - Yes, I had twins, triplets, etc. - No |  |
| Time of birth | At about what time was your child born? | Slider bar, labelled from left to right with:  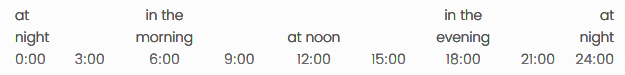 | |
| NICU transfer | Did your child require treatment in the neonatal intensive care unit (NICU) or the neonatology unit after birth? | - Yes - No |  |
| Nulliparous | Was this your first birth? | - Yes - No |  |
| Previous CS | Did you have a caesarean section for a previous birth? | - Yes - No |  |
| Place of birth | Where did you give birth? | - University hospital - Cantonal hospital - Regional hospital - Private hospital - Birthing centre - At home - Other: ____________ | “University hospital”, “Cantonal hospital” and “Regional hospital” were combined to “Public hospital” for statistical analyses. |
| Planned place of birth | You specified that you gave birth at [previous answer]. Is this where you were planning to give birth? | - Yes - No, the original plan was to give birth at: ________ |  |
| Known care provider | Did any of the health professionals who looked after you during labor also care for you during your pregnancy? | - Yes - No |  |
| Accompaniment | Who accompanied you at this birth? Multiple answers are possible. | - Husband/life partner - Family member - Doula - Friend - Nobody - Other: ____________ | As 98% of women were accompanied by their husband/life partner, the variation in this variable was low and it was therefore excluded from statistical analyses. |
| Preference for shared decision making | From your point of view, who should have made important decisions about how to proceed during childbirth?   - I should have made the decision on my own after I had been fully informed. - The health professional should have made the decision on my behalf after I had been fully informed. - I should have made the decision together with the health professional after I had been fully informed. | Likert scale:   - Strongly agree - Agree - Neither agree nor disagree - Disagree - Strongly disagree | Optimal scaling procedure and subsequent PCA. |
| Atmosphere | To what extent would you agree with the following statements regarding the birth?   - I felt that the health professionals were open to my wishes and needs. - The health professionals and I made the important decisions together. - I felt that the midwives and doctors worked well together. | Likert scale:   - Strongly agree - Agree - Neither agree nor disagree - Disagree - Strongly disagree | Chi-square tests between these single items and the item “pressure” revealed maximal odds ratios of 17.3 [95% CI 13.1, 22.72], suggesting that the variable “atmosphere” serves as a proxy rather than a predictor for pressure. It was therefore excluded from final analyses. |
| Birth mode preference | What kind of birth did you want **while you were pregnant**? | Visual analog scale, ranging from “Definitely a spontaneous vaginal delivery” (left) to “Definitely a caesarean section” (right) |  |
| Birth mode | How did you actually give birth? | - Spontaneous vaginal delivery - Forceps or vacuum delivery - Caesarean section |  |
|  | You stated that you had a caesarean section. **When** was it decided that you should have a caesarean section? | - The caesarean section was already planned before going to the hospital. - After delivery had already begun. - I required an emergency caesarean after less than 15 minutes. | Coded as “planned CS”, “unplanned CS” and “emergency CS” |
| Indication Planned CS | What was the reason you had a caesarean? Multiple answers are possible. | - I had health problems (e.g. preeclampsia). - Caesarean during a previous birth - Estimated weight of child at birth - Position/presentation of the baby (e.g. breech position) - Multiple birth (twins, triplets etc.) - The child was unwell. - It’s what I wanted. - I didn’t understand the reason. - I can’t remember. - Other reasons: _________________ |  |
| Reason elective CS | Why was it you wanted a caesarean section? Multiple answers are possible. | - Fear of pain or complications - Fear for my child’s safety - Earlier negative birth experience - Ability to plan the birth - To avoid injuries in the genital area - Other reasons: ________________ |  |
| Indication unplanned CS | What was the reason you had a caesarean? Multiple answers are possible. | - Health problems. - Prolonged labour/failure to progress - Position/presentation of the child - Failed induction of labour. - The child was unwell. - The pain was too severe. - I was exhausted. - I didn’t understand the reason. - I can’t remember. - Other reasons: _________________ |  |
| Indication instrumental vaginal birth | Why was it you had a forceps or vacuum delivery? Multiple answers are possible. | - The child was unwell. - Prolonged labour/failure to progress - I was exhausted. - I didn’t understand the reason. - I can’t remember. - Other reasons: _________________ |  |
| Duration of birth | **Roughly** how many hours were you in labour **after your contractions became regular**? | Slider bar, ranging from “0” (left) to “48+” (right). |  |
| Interventions | What (other) procedures were performed during the birth? Multiple answers are possible. | - Induction of labour - Vaginal examination - Electronic monitoring of contractions and heartbeat (CTG) - Episiotomy (surgical cut to enlarge opening of vagina) - Manual pressure on the abdomen to speed up birth - Rupture of membranes to release amniotic fluid (“breaking the water”) - I can’t remember. - None - Other procedures: _________________ |  |
| CTG | You indicated that monitoring of your child’s heartbeat and your contractions (CTG) was applied. Did you find the CTG uncomfortable? | - Yes - No |  |
| Vaginal examinations | Which of the following statements apply to the **vaginal examination(s)**?   - They did everything to make the examination bearable. - I was examined too often. - My privacy was respected. | Likert scale:   - Strongly agree - Agree - Neither agree nor disagree - Disagree - Strongly disagree |  |
| Indication induction of labor | Why was it that labour was induced in your case? Multiple answers are possible. | - The baby was unwell. - Gestational diabetes - I had other health problems (e.g. high blood pressure, preeclampsia). - Overdue/prolonged pregnancy - The child was considered too large. - Premature rupture of membranes and risk of infection - There were problems with the amniotic fluid (“the waters”). - My baby did not grow as expected. - I didn’t understand the reason. - I can’t remember. - Other reasons: _________________ |  |
| Indication episiotomy | Why was it you had an episiotomy? Multiple answers are possible. | - The child was unwell. - The baby was too big. - To avoid more severe injuries in the genital area. - Prolonged labour/failure to progress - I didn’t understand the reason. - I can’t remember. - Other reasons: _________________ |  |
| Indication amniotomy | Why was it you had induced rupture of your membranes? Multiple answers are possible. | - To speed up the birth - The child was unwell. - I didn’t understand the reason. - I can’t remember. - Other reasons: _________________ |  |
| Medication | Which medications or painkillers did you receive? Multiple answers are possible. | - Epidural anaesthesia (EDA, pain relief administered via the spine) - Patient-controlled button for analgesia (PCA pump) - Laughing gas - Other painkillers - Labour-inducing drugs - Labour-inhibiting drugs - Antibiotics - I receive some medication, but I’m not sure what it was. - None - Other: _________________ |  |
| Freedom of movement (CS) | Before you had to have a caesarean section, were you able to move around and choose the position you found most comfortable during the birth? | - Yes - No, because of the epidural anaesthesia (EDA) - No, because of the CTG (foetal monitoring). - No, for other reasons: _________________ |  |
| Freedom of movement (vaginal birth) | Were you able to move around and choose the position you found most comfortable during the birth? Multiple answers are possible. | - Yes - No, because of the epidural anaesthesia (EDA) - No, because of the CTG (foetal monitoring). - No, for other reasons: _________________ |  |
| Denied procedures | Were you **denied** any procedures (examinations, treatments) during the birth, even though you expressly requested them? | - No - Yes, namely: _________________ |  |
| Postnatal debriefing | After you gave birth, did you have the opportunity to discuss the birth with the health professionals involved? | - Yes - I didn’t feel the need. - No, because: _________________ |  |
| Postnatal debriefing helpful | Did this follow-up interview help you clear up or process something important? | - Yes - No |  |
| Maternal age | How old are you? | - 18-23 - 24-27 - 28-31 - 32-35 - 36-39 - 40+ |  |
| Nationality | What is your nationality? | - Swiss - German - French - Italian - Austrian - Afghan - Bosnian/Herzegovian - Brazilian - Chinese - Croatian - Eritrean - Hungarian - Kosovan - Macedonian - Polish - Portuguese - Romanian - Russian - Serbian - Slovakian - Spanish - Sri Lankan - Syrian - Turkish - US citizen - UK citizen - Other | “German”, “French”, “Italian” and “Austrian” were combined to “Neighboring state”, all other non-Swiss nationalities were combined to “other”. |
| Birth canton | In which canton was your child born? | - Aargau - Appenzell Ausserrhoden - Appenzell Innerrhoden - Basel-Landschaft - Basel-Stadt - Berne - Freiburg - Geneva - Glarus - Grisons - Jura - Lucerne - Neuenburg - Nidwalden - Obwalden - Schaffhausen - Schwyz - Solothurn - St. Gallen - Ticino - Thurgau - Uri - Vaud - Wallis - Zug - Zürich |  |
| Urbanization | Federal Statistical Office (FSO), [www.bfs.admin.ch](http://www.bfs.admin.ch) , Structure of the permanent resident population by canton, 1999-2018 |  | Based on the canton where the child was born (previous question), we used the percentage of permanent residents living in an “urban core area” or “area influenced by urban cores” as an indicator of urbanization within each canton. |
| Marital status | What is your marital status? | - Married/civil partnership - Single - Divorced - Widowed | Because <2% were either divorced or widowed, we combined them with the category “single”. |
| Socio-economic status | What is your net monthly household income? (the current income of all household members together) | - less than CHF 3,000 - CHF 3,000 – 4,999 - CHF 5,000 – 6,999 - CHF 7,000 – 8,999 - CHF 9,000 – 11,999 - CHF 12,000 – 15,000 - more than CHF 15,000 | Questions on income and mother’s education were reduced to socio-economic status using optimal scaling procedure and subsequent PCA. |
|  | What is your level of education or training? | - Compulsory education - Apprenticeship - Grammar school, vocational A level, specialized secondary school certificate (FMS), vocational college (DMS) - Higher technical and vocational training - University of Applied Sciences, educational college - University, Federal Institute of Technology (EPFL, ETH) - Other: __________________ |  |
| Health insurance | How are you insured? | - General - Semi-private - Private - I don’t know | Categories semi-private and private were combined to one category. |
