## Additional file 2: Table S2 for "Informal coercion during childbirth: risk factors and prevalence estimates from a nationwide survey among women in Switzerland"

Additional File 2: Table S2. Descriptive statistics of additional sociodemographic variables, pregnancy, and birth characteristics of the survey sample.

|  | **Survey** |
| --- | --- |
|  | ***N=6054*** |
| **Mother** | |
| Education: |  |
| Compulsory education | 129 (2.14%) |
| Apprenticeship | 1631 (27.1%) |
| Secondary school† | 445 (7.39%) |
| Higher technical and vocational training | 1075 (17.9%) |
| University of Applied Sciences, educational college | 1361 (22.6%) |
| University, ETHZ/EPFL | 1379 (22.9%) |
| Household income (CHF): |  |
| less than 3,000 | 242 (4.09%) |
| 3,000 – 4,999 | 665 (11.2%) |
| 5,000 – 6,999 | 1313 (22.2%) |
| 7,000 – 8,999 | 1497 (25.3%) |
| 9,000 – 11,999 | 1262 (21.3%) |
| 12,000 – 15,000 | 622 (10.5%) |
| more than 15,000 | 321 (5.42%) |
| Health insurance: |  |
| General | 4788 (79.8%) |
| Semi-private | 978 (16.3%) |
| Private | 234 (3.90%) |
| **Pregnancy** | |
| Gestational age (weeks): |  |
| < 32 | 31 (0.51%) |
| 32 - 36 | 329 (5.45%) |
| 37 - 41 | 4823 (79.9%) |
| > 41 | 853 (14.1%) |
| Parity: |  |
| Nulliparous | 3506 (57.9%) |
| Multiparous – no previous CS | 1926 (31.8%) |
| Multiparous – previous CS | 620 (10.2%) |
| Multiple birth | 79 (1.30%) |
| Risk pregnancy: |  |
| No | 4401 (72.8%) |
| Yes, with in-patient stay | 441 (7.29%) |
| Yes, with out-patient treatment | 1206 (19.9%) |
| Main caregiver: physician | 5145 (85.1%) |
| **Birth** | |
| Place of birth: |  |
| University hospital | 683 (11.3%) |
| Cantonal hospital | 2022 (33.6%) |
| Regional hospital | 1750 (29.1%) |
| Private hospital | 1002 (16.6%) |
| Birthing center | 338 (5.61%) |
| At home | 228 (3.79%) |
| Unplanned place of birth | 413 (6.83%) |
| Birth during business hours (7:00-17:00) | 2895 (47.8%) |
| Birth duration (h; SD) | 10.7 (9.96) |
| Mode of delivery: |  |
| Non-instrumental vaginal birth | 3952 (65.3%) |
| Forceps or vacuum birth | 693 (11.4%) |
| Planned cesarean section | 564 (9.32%) |
| Unplanned cesarean section | 505 (8.34%) |
| Emergency cesarean section | 340 (5.62%) |
| Induction of labor | 1533 (28.0%) |
| Episiotomy | 693 (12.6%) |
| Amniotomy | 1359 (24.8%) |
| Fundal pressure | 371 (6.77%) |
| Birth weight (g; SD) | 3343 (501) |
| Transfer to neonatal intensive care unit | 495 (8.18%) |

†Grammar school, vocational A level, specialized secondary school certificate (FMS), vocational college (DMS)
