## Additional file 3: Table S3 for "Informal coercion during childbirth: risk factors and prevalence estimates from a nationwide survey among women in Switzerland"

Additional File 3: Table S3. Pairwise associations between the experience of informal coercion and medical indications, other interventions, and diagnostic procedures.

|  |  | **Informal coercion** | |  |
| --- | --- | --- | --- | --- |
|  | ***n*** | **No** | **Yes** | ***p*** |
|  |  | ***N=4184*** | ***N=1300*** |  |
| **Indication for planned cesarean section** | | | | |
| Mother's health issues | 76 (13.3%) | 57 (12.9%) | 19 (14.8%) | 0.672 |
| Previous cesarean section | 197 (34.6%) | 156 (35.3%) | 41 (32.0%) | 0.563 |
| Child's weight | 71 (12.5%) | 53 (12.0%) | 18 (14.1%) | 0.636 |
| Child's position | 221 (38.8%) | 162 (36.7%) | 59 (46.1%) | 0.068 |
| Multiple birth | 29 (5.09%) | 21 (4.75%) | 8 (6.25%) | 0.652 |
| Child unwell | 16 (2.81%) | 12 (2.71%) | 4 (3.12%) | 0.765 |
| Mother's wish | 94 (16.5%) | 83 (18.8%) | 11 (8.59%) | 0.009 |
| Did not understand | 1 (0.18%) | 0 (0.00%) | 1 (0.78%) | 0.225 |
| **Reason for elective cesarean section** | | | | |
| Fear of pain/complications | 39 (41.5%) | 37 (44.6%) | 2 (18.2%) | 0.115 |
| Fear for child's safety | 49 (52.1%) | 43 (51.8%) | 6 (54.5%) | 1.000 |
| Previous negative birth experience | 25 (26.6%) | 23 (27.7%) | 2 (18.2%) | 0.721 |
| Ability to plan birth | 26 (27.7%) | 22 (26.5%) | 4 (36.4%) | 0.490 |
| Avoid genital injuries | 35 (37.2%) | 33 (39.8%) | 2 (18.2%) | 0.202 |
| **Indication for unplanned cesarean section** | | | | |
| Mother's health issues | 99 (11.8%) | 47 (9.33%) | 52 (15.5%) | 0.009 |
| Prolonged labor | 432 (51.5%) | 268 (53.2%) | 164 (49.0%) | 0.260 |
| Child's position | 366 (43.6%) | 232 (46.0%) | 134 (40.0%) | 0.098 |
| Failed induction of labor | 187 (22.3%) | 99 (19.6%) | 88 (26.3%) | 0.030 |
| Child unwell | 332 (39.6%) | 185 (36.7%) | 147 (43.9%) | 0.045 |
| Mother's pain | 53 (6.32%) | 33 (6.55%) | 20 (5.97%) | 0.848 |
| Mother exhausted | 103 (12.3%) | 62 (12.3%) | 41 (12.2%) | 1.000 |
| Did not understand | 14 (1.67%) | 3 (0.60%) | 11 (3.28%) | 0.007 |
| **Indication for forceps or vacuum birth** | | | | |
| Child unwell | 330 (47.6%) | 189 (44.9%) | 141 (51.8%) | 0.087 |
| Prolonged labor | 344 (49.6%) | 213 (50.6%) | 131 (48.2%) | 0.584 |
| Mother exhausted | 168 (24.2%) | 106 (25.2%) | 62 (22.8%) | 0.532 |
| Did not understand | 27 (3.90%) | 7 (1.66%) | 20 (7.35%) | <0.001 |
| **Indication for induction of labor** | | | | |
| Child unwell | 66 (4.32%) | 38 (4.04%) | 28 (4.75%) | 0.591 |
| Gestational diabetes | 149 (9.74%) | 86 (9.15%) | 63 (10.7%) | 0.366 |
| Mother's health issues | 198 (12.9%) | 125 (13.3%) | 73 (12.4%) | 0.664 |
| Overdue/prolonged pregnancy | 557 (36.4%) | 354 (37.7%) | 203 (34.5%) | 0.227 |
| Child too large | 122 (7.98%) | 66 (7.02%) | 56 (9.51%) | 0.099 |
| Rupture of membranes | 279 (18.2%) | 179 (19.0%) | 100 (17.0%) | 0.343 |
| Amniotic fluid issues | 189 (12.4%) | 113 (12.0%) | 76 (12.9%) | 0.667 |
| Child too small | 93 (6.08%) | 47 (5.00%) | 46 (7.81%) | 0.033 |
| Did not understand | 25 (1.64%) | 3 (0.32%) | 22 (3.74%) | <0.001 |
| **Indication for episiotomy** | | | | |
| Child unwell | 202 (29.1%) | 117 (28.7%) | 85 (29.7%) | 0.847 |
| Child too large | 125 (18.0%) | 89 (21.9%) | 36 (12.6%) | 0.002 |
| Avoid genital injuries | 210 (30.3%) | 132 (32.4%) | 78 (27.3%) | 0.170 |
| Prolonged labor | 220 (31.7%) | 141 (34.6%) | 79 (27.6%) | 0.061 |
| Did not understand | 63 (9.09%) | 10 (2.46%) | 53 (18.5%) | <0.001 |
| **Indication for amniotomy** | | | | |
| Speed up birth | 1022 (75.3%) | 718 (74.3%) | 304 (77.9%) | 0.174 |
| Child unwell | 66 (4.86%) | 44 (4.55%) | 22 (5.64%) | 0.480 |
| Did not understand | 55 (4.05%) | 22 (2.28%) | 33 (8.46%) | <0.001 |
| **Other interventions and diagnostic procedures** | | | | |
| Fundal pressure | 371 (6.77%) | 231 (5.52%) | 140 (10.8%) | <0.001 |
| Found CTG uncomfortable | 703 (16.2%) | 425 (13.2%) | 278 (25.0%) | <0.001 |
| Vaginal examinations (VE) were bearable: |  |  |  | <0.001 |
| Strongly agree | 2187 (59.0%) | 1826 (65.4%) | 361 (39.4%) |  |
| Agree | 1157 (31.2%) | 802 (28.7%) | 355 (38.8%) |  |
| Neither agree nor disagree | 228 (6.15%) | 124 (4.44%) | 104 (11.4%) |  |
| Disagree | 106 (2.86%) | 34 (1.22%) | 72 (7.86%) |  |
| Strongly disagree | 31 (0.84%) | 7 (0.25%) | 24 (2.62%) |  |
| VE were conducted too often: |  |  |  | <0.001 |
| Strongly agree | 118 (3.18%) | 47 (1.68%) | 71 (7.77%) |  |
| Agree | 242 (6.53%) | 115 (4.12%) | 127 (13.9%) |  |
| Neither agree nor disagree | 669 (18.1%) | 451 (16.2%) | 218 (23.9%) |  |
| Disagree | 1728 (46.6%) | 1354 (48.5%) | 374 (40.9%) |  |
| Strongly disagree | 949 (25.6%) | 825 (29.5%) | 124 (13.6%) |  |
| Privacy was respected during VE: |  |  |  | <0.001 |
| Strongly agree | 2055 (55.5%) | 1706 (61.1%) | 349 (38.1%) |  |
| Agree | 1271 (34.3%) | 890 (31.9%) | 381 (41.6%) |  |
| Neither agree nor disagree | 210 (5.67%) | 115 (4.12%) | 95 (10.4%) |  |
| Disagree | 120 (3.24%) | 55 (1.97%) | 65 (7.10%) |  |
| Strongly disagree | 49 (1.32%) | 24 (0.86%) | 25 (2.73%) |  |
| **Anesthesia, painkillers, and other medication** | | | | |
| Spinal/epidural anesthesia | 2311 (42.2%) | 1598 (38.2%) | 713 (54.9%) | <0.001 |
| PCA | 399 (7.29%) | 282 (6.75%) | 117 (9.01%) | 0.007 |
| Laughing gas | 781 (14.3%) | 566 (13.5%) | 215 (16.6%) | 0.008 |
| Labor-inducing drugs | 1654 (30.2%) | 1105 (26.4%) | 549 (42.3%) | <0.001 |
| Labor-inhibiting drugs | 590 (10.8%) | 399 (9.55%) | 191 (14.7%) | <0.001 |
| **Birth debriefing** | | | | |
| Opportunity to discuss birth afterwards: |  |  |  | <0.001 |
| Yes | 2709 (49.4%) | 2145 (51.3%) | 564 (43.4%) |  |
| No | 865 (15.8%) | 478 (11.4%) | 387 (29.8%) |  |
| No need | 1908 (34.8%) | 1559 (37.3%) | 349 (26.8%) |  |
| Debriefing was helpful | 2220 (82.5%) | 1804 (84.7%) | 416 (74.2%) | <0.001 |

Note: *n* indicates the number of women who were asked a specific question, which varied from item to item due to skip patterns. *p* values are derived from χ^2^ test or Fisher’s exact test.
